# Saliva testing is accurate for early-stage and presymptomatic COVID-19

**DOI:** 10.1101/2021.03.03.21252830

**Authors:** Abigail J. Johnson, Shannon Zhou, Suzie L. Hoops, Benjamin Hillmann, Matthew Schomaker, Robyn Kincaid, Jerry Daniel, Kenneth Beckman, Daryl M. Gohl, Sophia Yohe, Dan Knights, Andrew C. Nelson

**Affiliations:** Division of Epidemiology and Community Health, School of Public Health, University of Minnesota; BioTechnology Institute, College of Biological Sciences, University of Minnesota; Department of Computer Science and Engineering, University of Minnesota; M Health Fairview, Minneapolis; University of Minnesota Genomics Center, University of Minnesota; Department of Laboratory Medicine and Pathology, Division of Molecular Pathology and Genomics, University of Minnesota; Department of Genetics, Cell Biology, and Development, University of Minnesota

**Author notes:** For correspondence: Andrew C. Nelson, Department of Laboratory Medicine and Pathology, Division of Molecular Pathology and Genomics, University of Minnesota, 420 Delaware St. S.E., MMC 609, Minneapolis, MN 55455. Andrew C. Nelson and Dan Knights contributed equally to this manuscript.

## Abstract

**Background:** Although nasopharyngeal (NP) samples have been considered the gold standard for COVID-19 testing, variability in viral load across different anatomical sites could theoretically cause NP samples to be less sensitive than saliva or nasal samples in certain cases. Self-collected samples also have logistical advantages over NP samples, making them amenable to population-scale screening.

**Methods:** To evaluate sampling alternatives for population screening, we collected NP, saliva, and nasal samples from two cohorts with varied levels and types of symptoms.

**Results:** In a mixed cohort of 60 symptomatic and asymptomatic participants, we found that saliva had 88% concordance with NP when tested in the same testing lab (n = 41), and 68% concordance when tested in different testing labs (n = 19). In a second cohort of 20 participants hospitalized for COVID-19, saliva had 74% concordance with NP tested in the same testing lab, but detected virus in two participants that tested negative with NP on the same day. Medical record review showed that the saliva-based testing sensitivity was related to the timing of symptom onset and disease stage.

**Conclusions:** We find that no sample site will be perfectly sensitive for COVID-19 testing in all situations, and the significance of negative results will always need to be determined in the context of clinical signs and symptoms. Saliva retained high clinical sensitivity while allowing easier collection, minimizing the exposure of healthcare workers and need for personal protective equipment, and making it a viable option for population-scale testing.

## Introduction

Throughout the COVID-19 pandemic, the spread of infection has significantly outpaced laboratory testing to identify SARS-CoV-2. Seroprevalence studies performed in March-May of 2020 in the United States suggested that the number of infections were perhaps 10-fold greater than confirmed laboratory diagnoses^1^. This scenario significantly challenged public health efforts to monitor and contain the spread of disease. Nasopharyngeal (NP) samples have been considered the gold standard for COVID-19 testing. However, alternative samples like self-collected saliva offer advantages for population-scale screening and may perform well in specific clinical situations.

A number of studies comparing saliva, oral, and/or nasal samples with NP samples have reported heterogeneity in sensitivity or positive percent agreement (PPA) ranging from 66%-98%^2–12^; this heterogeneous performance is likely impacted by differences in patient populations and methods of sample collection and processing. For example, optimization of saliva sample processing within one institution improved the performance of this sample type across two sequential studies ^3,5^. More importantly, clinical test performance is dependent on pre-analytic variables such as collection timing relative to the patient’s disease course and anatomic site of collection. A study of inpatients at an advanced disease stage showed that lower respiratory samples (bronchoalveolar lavage) were more frequently positive (93%) than pharyngeal (32%) or nasal (63%) samples^13^. Further studies of samples from different anatomic sites at different points in disease course are necessary to better understand how these variables impact clinical test performance.

Here, we acquired patient-collected saliva and anterior nasal research specimens for comparison with provider-collected NP samples in both outpatient and inpatient settings. The clinical context of specimen collection, the timing of sample collection during disease course, and the analytical performance of different molecular tests were assessed for their impact on the test result agreement between different anatomical sites.

## Methods

### Study design and cohort definition

Cohort 1 study participants, presenting with both symptomatic and asymptomatic concerns for COVID-19 were enrolled opportunistically from a population receiving a nasopharyngeal (NP) COVID-19 test in outpatient screening or emergency department (ED) settings. Due to limited testing resources available during the study, we relied on NP results from routine clinical testing. Nasal and saliva samples were prospectively collected and bio-banked for retrospective testing. After 30 positive cases were detected by clinical NP, enrollment was stopped and 30 negative cases were randomly selected for comparison.

Due to clinical test triage patterns outside the study’s purview, clinical NP samples were routed to different test platforms. We divided Cohort 1 into two groups to reflect testing methods used for the clinical NP test. Cohort 1A (outpatient) had clinical NP samples and bio-banked saliva and nasal samples tested on our Clinical Laboratory Improvement Amendments-certified Laboratory Developed Test (CLIA-LDT). Cohort 1B (ED) had clinical NP sample testing performed on one of two different commercial testing platforms.

Cohort 2 participants were recruited at admission from a population of hospitalized patients who had previously tested positive for COVID-19. In this group, NP, saliva, and nasal samples were obtained from participants for concurrent analysis on the CLIA-LDT. Both cohorts were recruited, consented, and enrolled under protocols approved by the University of Minnesota’s Institutional Review Board (Cohort 1A and 1B: STUDY00009393, Cohort 2: STUDY00009560).

### Sample collection, handling, and biobanking

NP samples were collected by a healthcare provider for all participants; saliva and anterior nasal samples were patient-collected under direct observation. Study participants were given saliva testing kits (Item OM-505, DNA Genotek, Ottawa, Ontario, Canada) and anterior nasal testing kits (Item OCD-100, DNA Genotek, Ottawa, Ontario, Canada) with written and verbal instructions from monitoring healthcare workers.

In Cohort 1, liquid saliva samples mixed with buffer and nasal swabs in buffer were transported at room temperature and stored at −20C, according to instructions from the manufacturer. Selected research samples were thawed at room temperature immediately prior to analysis on the CLIA-LDT. In Cohort 1, NP swabs were collected, handled, and processed immediately according to the normal course of clinical testing through the healthcare system (1A = CLIA-LDT; 1B = commercial assays).

In Cohort 2, all samples were collected simultaneously within 48 hours of admission. The three sample types were transported together at room temperature to the CLIA-LDT testing facility, and processed within 24 hours.

### Sample Extraction and Molecular Testing

A reverse transcription polymerase chain reaction (RT-PCR) based on primer-probe sets for the SARS-CoV-2 N gene (N1 and N2 targets) and human control ribonuclease P (RP) published by the United States Centers for Disease Control was validated for clinical use^14^ following the regulatory requirements of CLIA and the Federal Drug Administration’s Emergency Use Authorization criteria. A sample was reported positive for SARS-CoV-2 if either N1 or N2 viral targets were detected with Ct < 40 passing data quality control. Negative samples required the internal RP control was detected with Ct < 38. For saliva and nasal samples obtained in sample buffer, nucleic acid extraction was performed with the Promega Maxwell RSC Viral Total Nucleic Acid kit (Cat# AS1330, Promega, Madison, WI) on the Maxwell RSC instrument per manufacturer’s instructions. For Cohort 1B, NP swab sample testing was performed on the Cepheid Xpert Xpress SARS-CoV-2 test and the Diasorin Simplexa COVID-19 Direct assay following manufacturer’s instructions.

### Medical record review and symptom scoring

Medical records from telehealth, clinic, or hospital visits were reviewed for relevant symptoms including: loss of taste or smell, shortness of breath, cough, sore throat, fatigue, diarrhea, nausea or vomiting, loss of appetite, chest pain, and myalgia or headache. Physician and nursing notes immediately prior to the initial testing date and over the potentially symptomatic period (approximately 10-20 days after testing positive) were reviewed. Subjective symptoms were coded as either present (“Yes”) or absent (“No”) or as (“Mild”, “Moderate”, or “Severe”) when the medical record stated severity. In cases where an individual reported at least one symptom; the symptoms that were not reported were considered absent (“No”). In cases where there was no report of symptoms; symptoms were coded as “NA”. Objective signs of interest were defined as elevated body temperature (fever) and decreased oxygen saturation based on documentation in the medical record. Data abstraction from the medical records was completed by one study author and reviewed for accuracy by a second author.

### Statistical analysis

Group mean variation between NP, saliva, and nasal samples were assessed with the Kruskal-Wallis rank sum test. Correlations between methods were assessed using Pearson correlation coefficient. Bland-Altman analysis was used to assess method agreement between saliva or nasal samples relative to NP. PPA was calculated as the proportion of comparative method positives where the test method was positive. Overall percent agreement (OPA) was calculated as the proportion of tests where the test and comparative method agreed. To generate a symptom heat-map, symptoms were re-coded following the COVID-19 probability score (P(COVID)) prediction equation by Menni et al.^15^. Mild cough was re-coded as “No” and only severe fatigue was coded as “Yes”. Heat map rows are clustered by similarity using the complete linkage method and columns are sorted by cohort, method concordance, P(COVID), age, and sex. Average cycle threshold (Ct) was calculated for NP, saliva, and nasal samples as the mean Ct for N1 and N2. Relative viral load was calculated as ((2^(RP-N1)) + (2^(RP-N2)))/2. Data analysis and data visualization was completed using R version 3.4.3^16^ and the following packages: ggplot2 version 3.2.1^17^; cowplot version 0.9.4^18^; tidyverse version 1.3.0^19^; reshape 2 version 1.4.3^20^; pheatmap version 1.0.12^21^; viridis version 0.5.1^22^; RColorBrewer version 1.1-2^23^; and ggpubr 0.2.4^24^.

## Results

### Cohort Characteristics

Two distinct patient cohorts were included in the study (Figure 1). Cohort 1 consisted of 354 patients with clinical NP results from tests in outpatient or ED settings. Analysis per design was performed on 30 positive and 30 negative samples. The cohort was split into 1A (n=41) and 1B (n=19) based on heterogeneity of clinical NP test routing. Cohort 2 consisted of 20 inpatients from a dedicated COVID-19 ward for disease management; all had prior laboratory evidence of SARS-CoV-2 on clinical testing, but the timing of those results prior to admission was variable (1 day to 4 weeks).

**Figure 1:**
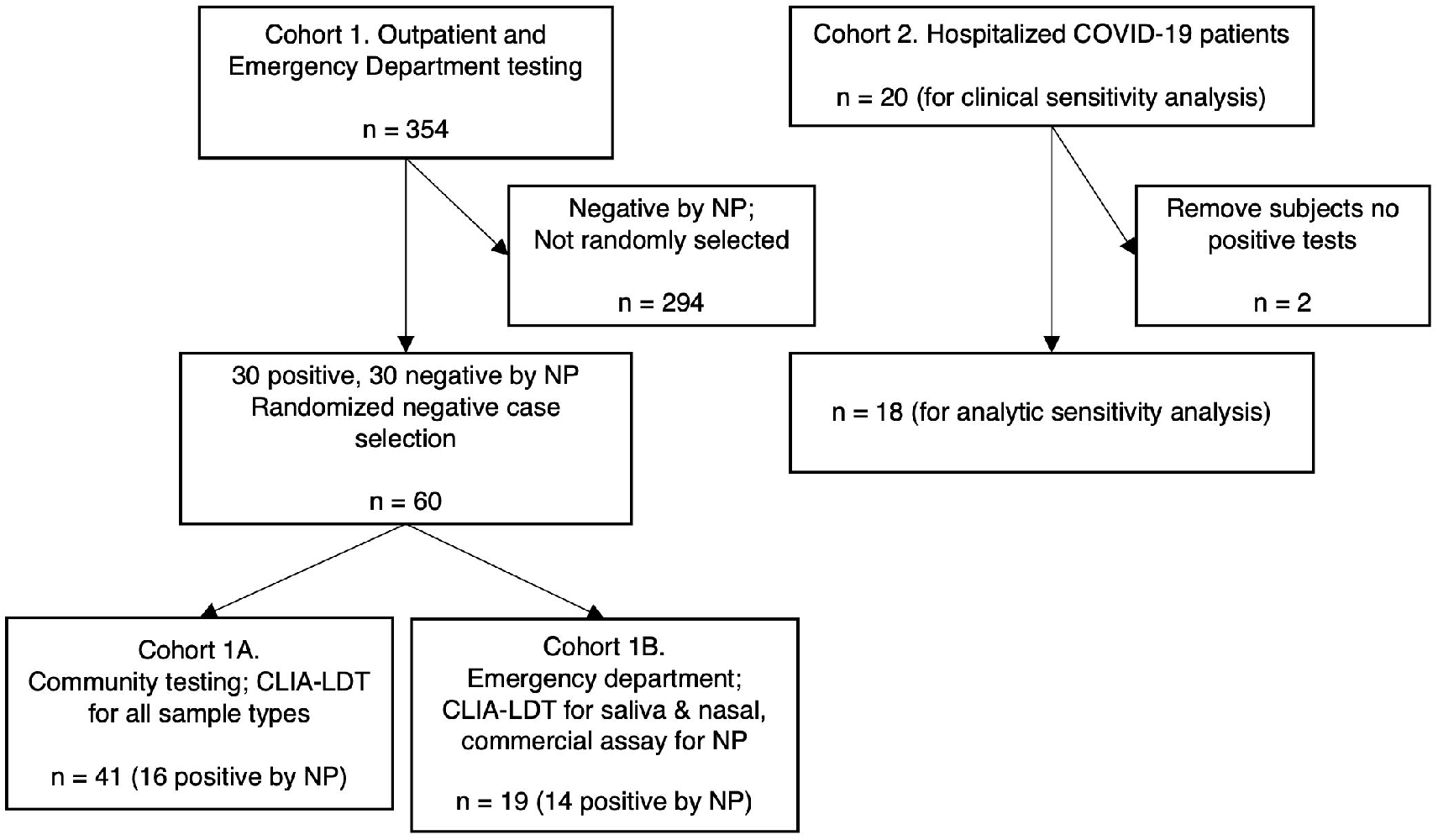
Cohort flow diagram. Cohort 1 was enrolled prospectively at outpatient screening and emergency department locations for sample collection and biobanking. When the pre-specified 30 positive samples were acquired, retrospective testing of saliva and anterior nasal samples was performed for 30 positives and 30 randomly selected negative samples. Heterogeneity in clinical NP test routing during the pandemic response necessitated sub-cohort analysis of 1A and 1B. Cohort 2 was enrolled and tested prospectively at an inpatient ward dedicated to COVID-19 patient care. Clinical sensitivity analysis is performed on all 20 patients enrolled in Cohort 2, while 2 patients were excluded from analytic sensitivity analysis due to negative test results for all three anatomic sites.

Cohort 1A had an average age of 39.6 years, and was skewed toward female gender at 92.7% Cohort 1B had an average age of 44.5 years, and was skewed toward male gender at 63.2%. Symptomatic patients comprised approximately 80% of both Cohorts 1A and 1B, although a higher proportion of symptomatic patients in Cohort 1B had positive clinical test results (92.3%) versus 1A (64.3%). Cohort 2 had an average age of 62.5 years, consistent with the increased hospitalization rate of older COVID-19 patients. Gender was equally distributed in Cohort 2. (Table 1).

**Table 1.**

| Characteristic | Cohort 1A | Cohort 1B | Cohort 2 |
| --- | --- | --- | --- |
| n | 41 | 19 | 20 |
| Age (years) | 39.6 ± 10.9 | 44.5 ± 20.1 | 62.5 ± 18.1 |
| Sex (% female) | 92.7% | 36.8% | 50.0% |
| % participants with symptoms | 77.8% | 81.3% | 100.0% |
| % participants with symptoms who are positive | 64.3% | 92.3% | 90.0% |
| % participants without symptoms who are positive | 0.0% | 33.3% | 0.0% |
| N1_NP tests with Ct values (n) | 16 | 14 | 16 |
| N1_Nasal tests with Ct values (n) | 14 | 8 | 11 |
| N1_Saliva tests with Ct values (n) | 13 | 8 | 12 |
| N2_NP tests with Ct values (n) | 16 | 14 | 16 |
| N2_Nasal tests with Ct values (n) | 13 | 7 | 11 |
| N2_Saliva tests with Ct values (n) | 13 | 8 | 14 |

N1 and N2 refer to primer-probe sets for the SARS-CoV-2 N gene (N1 and N2 targets). NP: Nasal pharyngeal.

### Concordance of Testing on Saliva, Nasal, and NP Samples

Clinical NP testing results in Cohort 1A (n=41) identified 16 positive and 25 negative patients. Banked saliva and nasal samples showed 87.5% PPA and 95.1% OPA, respectively, compared to NP (Table 2 and Supplemental Table 1). Saliva and nasal results were 100% concordant in Cohort 1A.

**Table 2.**
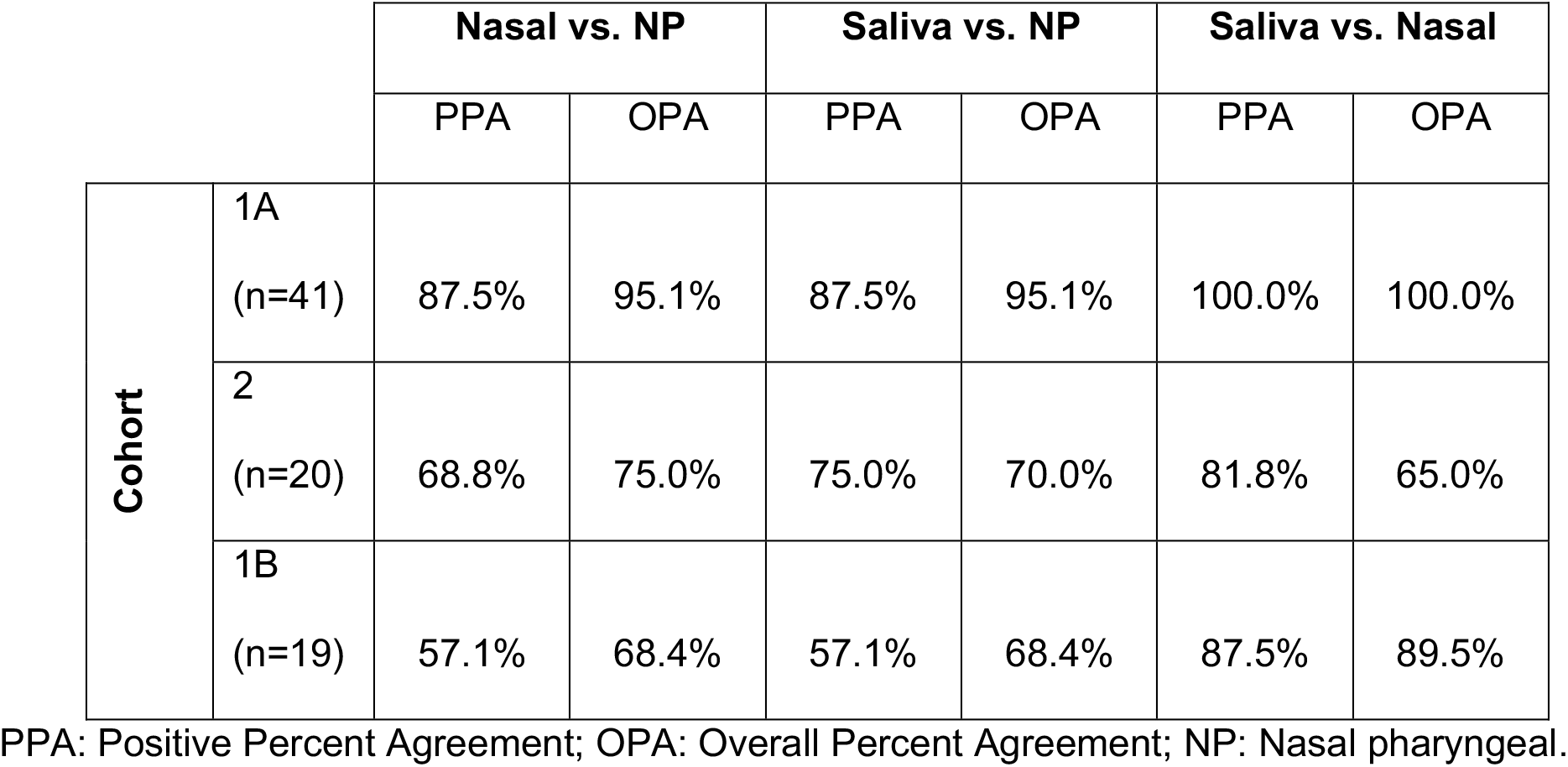

|  |  | Nasal vs. NP |  | Saliva vs. NP |  | Saliva vs. Nasal |  |
| --- | --- | --- | --- | --- | --- | --- | --- |
|  |  | PPA | OPA | PPA | OPA | PPA | OPA |
| **Cohort** | 1A<br>(n=41) | 87.5% | 95.1% | 87.5% | 95.1% | 100.0% | 100.0% |
|  | 2<br>(n=20) | 68.8% | 75.0% | 75.0% | 70.0% | 81.8% | 65.0% |
|  | 1B<br>(n=19) | 57.1% | 68.4% | 57.1% | 68.4% | 87.5% | 89.5% |

PPA: Positive Percent Agreement; OPA: Overall Percent Agreement; NP: Nasal pharyngeal.

Clinical NP testing on different commercial platforms in Cohort 1B (n=19) produced 14 positive and 5 negative results. Testing banked saliva and nasal samples on the CLIA-LDT provided more discordant results: the PPA for both saliva and nasal samples compared against NP was 57.1%. Further, there were two discrepancies between nasal and saliva sample results producing 87.5% PPA and 89.5% OPA between these sample types (Supplemental Table 2).

Simultaneous collection and processing of all samples for Cohort 2 (n=20) demonstrated 16 positive NP, 14 positive saliva, and 11 positive nasal samples. The PPA ranged from 69%-82% and the OPA from 65%-75% for all pairwise comparisons (Table 2). Saliva sampling identified 2 positive patients who tested negative by NP, and inversely NP testing identified 4 positive patients who tested negative by saliva (Supplemental Table 3). Nasal samples performed poorly in this patient cohort.

Clinical sensitivity in Cohort 2 was calculated on the full cohort with clinical COVID-19 diagnosis (n=20) showing: NP = 80%, saliva = 70%, and nasal = 55%. Eighteen patients (90%) were positive for SARS-CoV-2 by at least one upper respiratory sample type. Therefore, analytical sensitivity was calculated using this number as a denominator, showing: NP = 89%, saliva = 78%, and nasal = 61%.

### Comparison of Cycle Threshold Results Across Sample Type

Cycle threshold (Ct) values for each anatomic sample site were compared in Cohorts 1A and 2, in which all samples were analyzed on the same analytical platform. The interquartile ranges for both N1 and N2 were largely overlapping (Figure 2A, Supplemental Figure 1A). In Cohort 1A, the 2 discordant NP sample data points had Ct values of 36.6 and 38.4, near the assay limit of detection. Cohort 2 had more discordant data points, consistent with the higher average Ct value (lower relative viral load) observed in all anatomic sample types in this inpatient cohort tested at later disease stages. Discordant data points in this cohort ranged as low as 26.2, suggesting greater biologic variability in relative viral load between anatomic sites in patients at later disease stages. Similar results were observed with relative viral load (Supplemental Figure 2).

**Figure 2:**
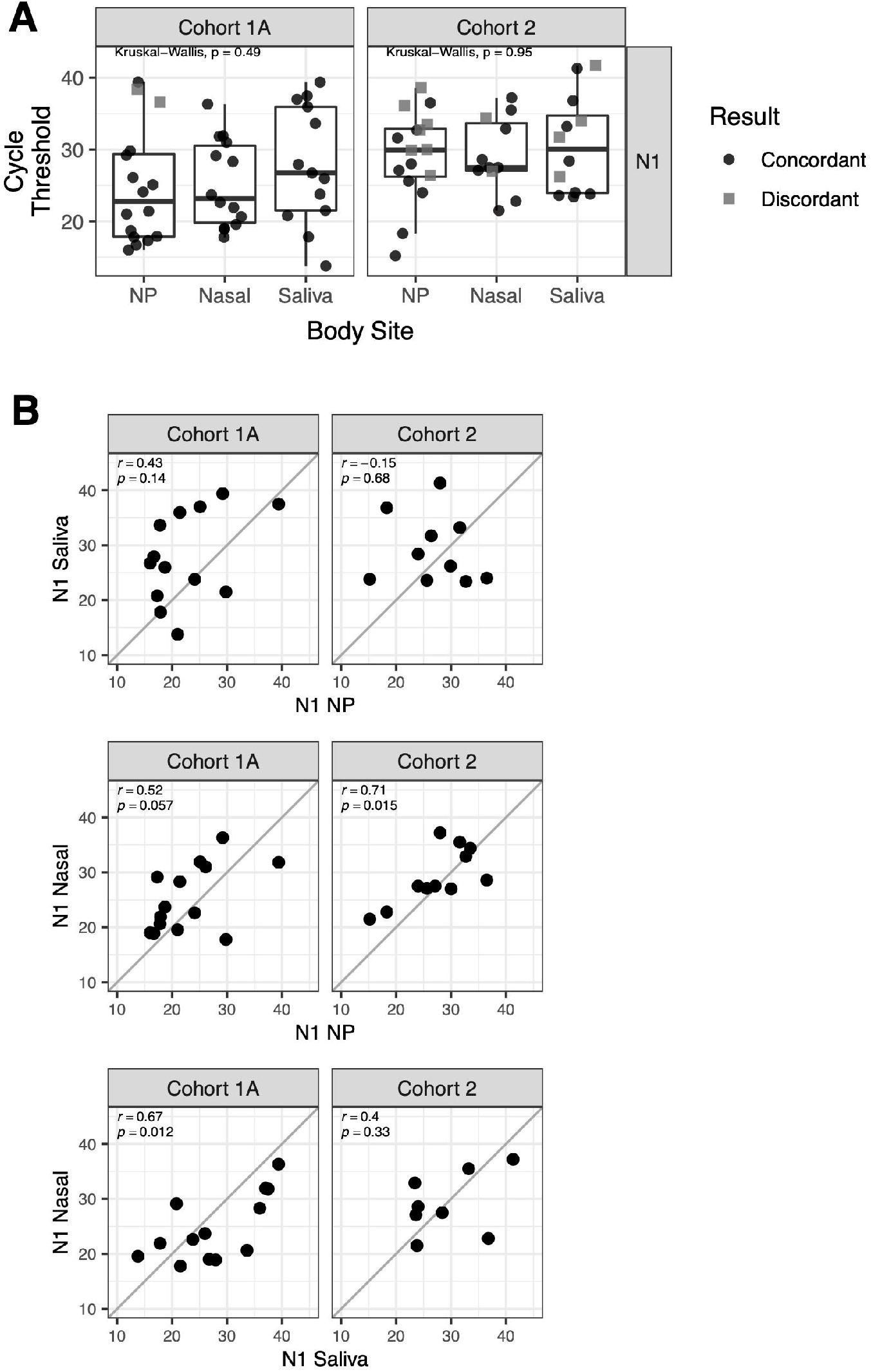
Cycle thresholds are similar across anatomic sites and cycle threshold agreement varies by anatomic site and cohort. **A)** Box plots showing cycle threshold (Ct) values for Cohort 1A (n = 41) and 2 (n = 18) from the CLIA-LDT assay for nasal pharyngeal samples (NP), nasal samples, and saliva samples using N1 primers. Black circles are samples with concordance between nasal or saliva samples and NP samples (positive/positive or negative/negative). Grey squares show discordance between nasal or saliva samples and NP samples (positive/negative). Groups are compared using Kruskal-Wallis test by ranks. **B)** Scatter plots of Ct values from N1 primers for concordant samples from different sample types: saliva v. NP; nasal v. NP; nasal v. saliva. Correlation was assessed using Pearson’s correlation. The correlation coefficient (r) and p-value for each comparison are indicated on each plot. See Supplemental Figure 1 for analogous analysis of N2 primers and Supplemental Figure 2 for analysis of relative viral load.

Comparison of N1 (Figure 2B) and N2 (Supplemental Figure 1B) Ct values between sample types from the same patient showed that saliva samples were heterogeneous in comparison to both NP and nasal samples, generally showing low, non-significant correlation. The correlation of NP vs. nasal Ct values was relatively tighter, with three of four comparisons trending towards or reaching statistical significance.

Bland-Altmann analysis using samples from Cohort 1A and Cohort 2 demonstrated a mean bias of −3.98 for N1 target Ct values and −5.3 for N2 target Ct values indicating average lower Ct values for NP samples compared to saliva samples (Figure 3A, B). Comparison of nasal and NP samples demonstrated a smaller mean bias of −2.07 for N1 and −3.73 for N2, again showing average lower Ct values for NP samples (Figure 3C, D). Heterogeneity around the mean bias was observed with relatively large limits of agreement (95% confidence ranges). There was no significant skewing or proportional difference at early or later Ct values.

**Figure 3:**
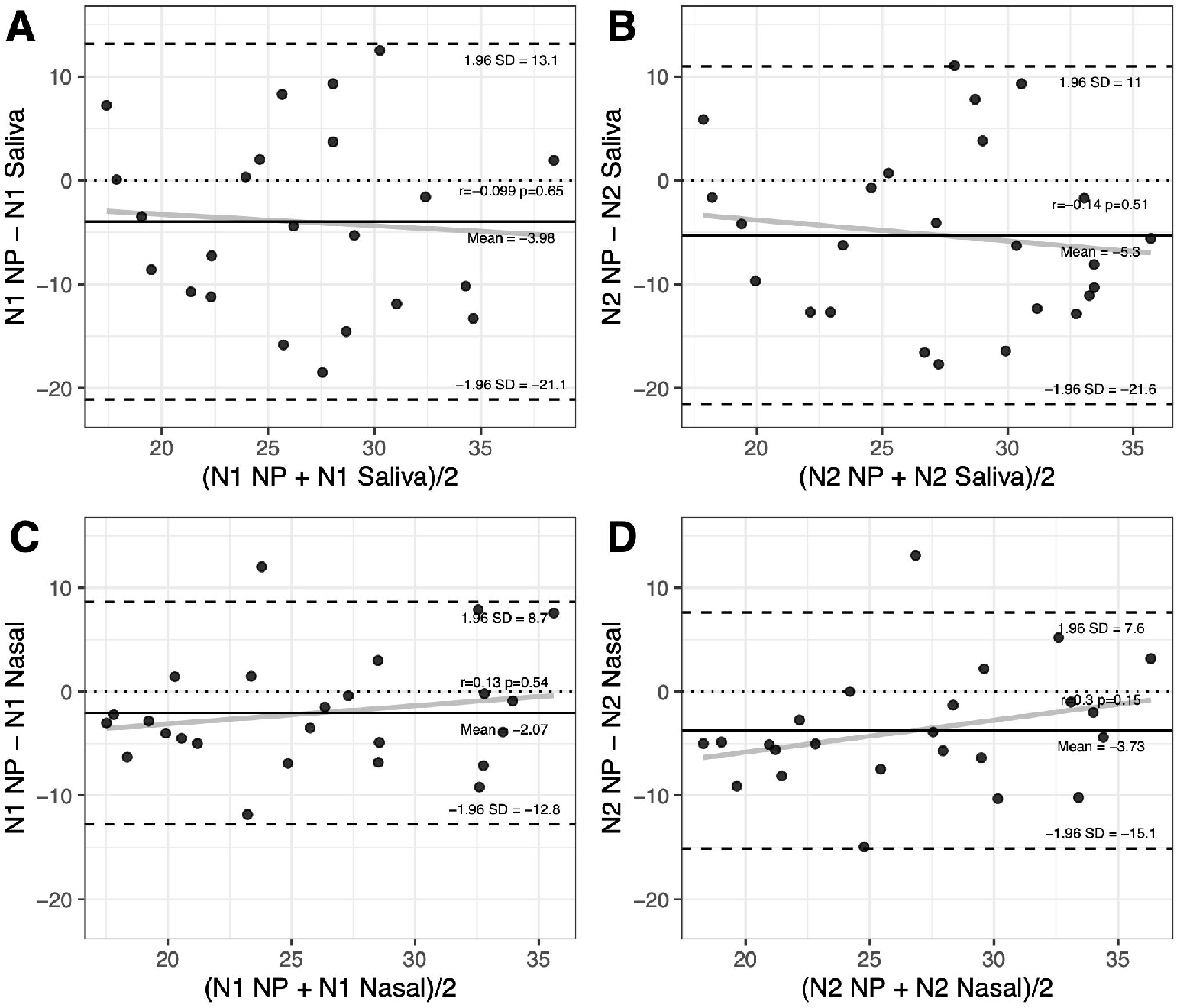
Bland-Altman analysis reveals minimal proportional bias between saliva and nasal cycle thresholds relative to NP. Bland-Altman (mean-difference) plots showing the relationship between average cycle threshold (Ct) and the difference between Cts for saliva v. nasal pharyngeal samples (NP) for **A)** N1 and **B)** N2 primers; and nasal v. NP for **C)** N1 and **D)** N2 primers. The solid black line shows the mean bias for each comparison. Dashed lines represent the limits of agreement (95% confidence interval) around the mean bias (+/-1.96*standard deviation [SD]). Solid grey line shows the linear relationship between the mean and difference. Pearson’s correlation (r) and p-value (p) is reported for the correlation between the mean and the difference.

### Correlation of Patient Symptoms with Test Results

Symptoms associated with COVID-19 were recorded from the medical record, scored, and used to calculate P(COVID) according to the Menni et al. prediction model^15^. Adequate records to calculate P(COVID) were available for 17 of 41 patients in Cohort 1A (outpatient), 16 of 19 patients in Cohort 1B (ED), and 19 of 20 patients in Cohort 2 (inpatient). Two patients in Cohort 1A had elevated P(COVID) scores (>0.5) but false negative saliva and nasal samples (Figure 4). Interestingly, both were being re-tested due to persistent symptoms at 2 and 4 weeks, respectively, after onset of their laboratory-confirmed COVID-19. No clear pattern of symptoms within Cohort 2 was evident in relation to the concordant or discordant test results from the three anatomical sites (Figure 4). In Cohort 1B, discrepancies between elevated P(COVID) scores and negative saliva tests (Supplemental Figure 3A) occurred in three patients without objective fever or oxygen saturation abnormalities who had complete resolution of subjective symptoms in <48 hours. We note that the P(COVID) score is heavily weighted toward loss of taste or smell, a subjective symptom. Saliva and nasal samples demonstrated complete clinical agreement with NP samples in patients with low P(COVID) scores (<0.5) who were tested when initial symptoms developed; suggesting that saliva and nasal samples perform well in the population screening setting.

**Figure 4:**
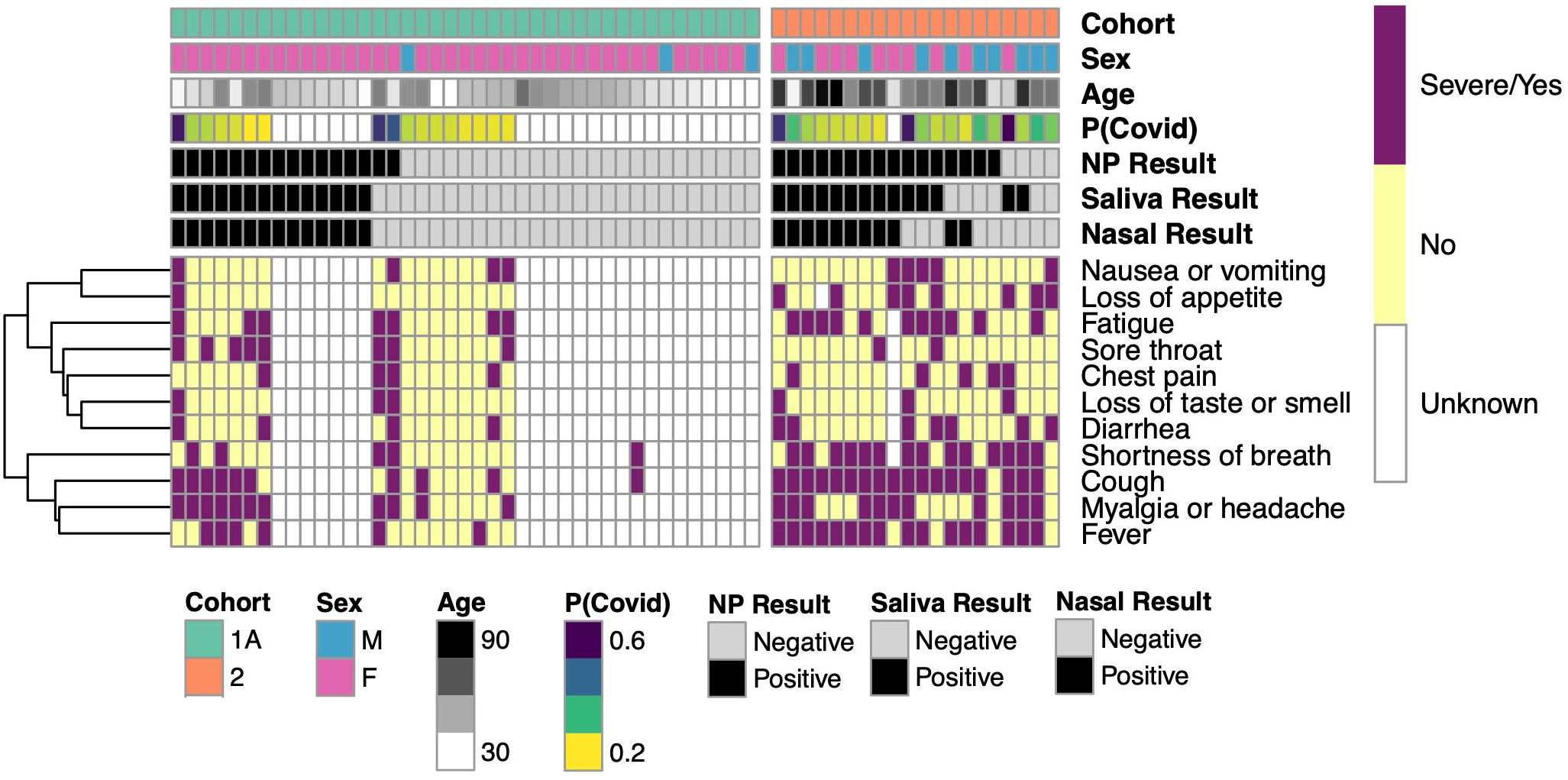
Symptom heat-map for agreement between positive and negative test results by test method and cohort. Heat map columns are participants and rows are symptoms. Symptom presence is indicated as either severe or yes (purple), no (yellow), or unknown (white). Symptoms are clustered by similarity and clustering is indicated by the dendrogram (left). Test results are indicated by color as either positive (black) or negative (light grey). Each participant’s calculated probability of COVID-19 (P(COVID)) is indicated with higher probabilities shown in purple and lower probabilities in yellow. Participant sex, age, and cohort are also indicated.

## Discussion

COVID-19 is a challenge to diagnose and treat at the individual patient level and to manage through public health measures at the community level. Alternative sample types such as patient-acquired saliva and anterior nasal samples are needed to facilitate high-volume testing when there are shortages of qualified healthcare workers and necessary supplies to support provider-acquired NP sampling. Our study sought to compare the performance of saliva and anterior nasal samples with standard of care NP samples for SARS-CoV-2 detection in different clinical settings. We observed variability in test results dependent on the clinical setting where samples were collected and the heterogeneous pattern of clinical test routing that was an uncontrolled variable in Cohort 1. Our results highlight some of the challenges facing diagnostic characterization of COVID-19 and indicate good performance of saliva and anterior nasal samples to detect SARS-CoV-2 virus when patients present for outpatient testing early in their disease course.

Variability in laboratory test results is caused by both pre-analytical and analytical factors. COVID-19 is a biologically heterogeneous infection: the anatomic distribution of active viral replication and the severity of symptoms in comparison to viral load are variable between different patients and during each individual’s disease course. These are important pre-analytical factors impacting test performance on samples from different anatomical sites. A recent systematic review and meta-analysis of saliva samples found that the pooled sensitivity of saliva (83%) was similar to NP swab (84%) with diagnostic equivalency highest in the ambulatory setting^25^. Additionally, recent pre-print data has been released supporting the hypothesis that saliva viral load may be a strong clinical predictor of COVID-19 severity^26^. These data overall support the important role saliva testing can play in assessing COVID-19.

We observed good PPA in Cohort 1A between the testing of banked saliva and nasal samples compared to the prior clinical NP result. Analytical variation was minimized in this cohort, with all samples tested using the same RT-PCR assay. Saliva and nasal samples showed complete agreement, and only two patients (of 16 total positive) had false negative saliva and nasal sample results compared to NP. These patients had previous laboratory-confirmed COVID-19 infections and were being re-tested due to persistent symptoms. The clinical NP sample for both of these patients had Ct values consistent with viral loads below the 95% confidence limit of detection for the clinical assay. Therefore, the false negative results in these cases could be due to degradation of the low viral load during the freeze-thaw cycle inherent to the pre-analytical study sample handling for this cohort. Considering only patients presenting within the first 10 days of symptom onset (n=14), saliva, nasal and NP samples had 100% PPA, suggesting that all three anatomical sample sites perform well for initial diagnosis of COVID-19.

Biologic heterogeneity regarding persistence and anatomic distribution of viral replication later in COVID-19 course was apparent in Cohort 2 (inpatient setting). An analytical advantage of Cohort 2 was the prospective, parallel collection of all three sample types with simultaneous handling and testing on the same platform. In this setting, no single anatomical site provided a positive result for all 18 patients with at least one positive sample type. NP and saliva samples performed better than nasal samples; however, the combination of an observed, patient-collected nasal and saliva sample together detected the same number of unique positive patients (n=16) as did the provider-collected NP sample. Theoretically, combining patient-collected saliva and nasal samples into the same collection-stabilization buffer could improve clinical sensitivity. Finally, there were 9 of 18 positive patients lacking SARS-CoV-2 detection in at least one of the three anatomical sites, and there were 4 patients with only 1 of 3 sites testing positive. This heterogeneity suggests that seeking a negative COVID-19 molecular test to guide further patient management has limited clinical value, as a negative test from one anatomical site does not rule out the presence of viral RNA in other sites from the same patient.

The data from Cohort 1B are difficult to interpret confidently. The unintended use of different molecular tests for the clinical NP test was an analytical confounder. Though the limit of detection for the commercial platforms (250-500 viral copies/mL) is lower than the CLIA-LDT (1670 copies/mL), independent clinical quality assurance data from our laboratory (see Supplemental Materials) comparing replicate testing of NP samples between platforms showed PPA of 90-97%, a level of agreement aligned with published data using standard of care NP samples^27–29^. This suggests additional pre-analytical variables impacted the poor concordance observed in Cohort 1B. The clinical vignettes of discrepant results across anatomical sites and testing platforms in Cohort 1B highlight the challenges of medical decision making with limited knowledge of the associations between laboratory test data and the natural biology of SARS-CoV-2 infection.

Our results highlight the importance of clinical judgement and integration of the patient’s medical presentation when interpreting individual test results. Understanding how anatomic site, timing of collection in disease course, handling and transport of the specimen, and analytical platform can influence test results is crucially important to making informed medical decisions regarding COVID-19 management^30^. Continued study of these factors in diverse clinical settings is necessary for the medical field to improve the response to this global pandemic. Overall, our findings support the conclusion that self-collected saliva testing is effective for COVID-19 detection, especially in early stages of disease progression.

## Data Availability

Deidentified data is available on request.

## Acknowledgements

The authors thank Beth Jorgenson, Sandra Tekmen, Krista Goldsmith, Andrew Snyder, Stephanie McGlone, and Jill Cordes for their efforts in coordination of sample collection. We thank Drs. Tyler Bold, Peter Southern and their laboratory staff for sample management and safety protocols. We want to acknowledge the significant efforts towards the COVID-19 testing effort made by faculty and staff at: the University of Minnesota Genomics Center (Benjamin Auch, Dr. Patrick Grady, Darrell Johnson, Ray Watson, Lindsey Gengelbach, Dinesha Walek, Paige Marsolek, Shea Anderson and many others), the Department of Laboratory Medicine & Pathology (Drs. Leo Furcht, Patricia Ferrieri, Anthony Killeen, Bharat Thyagarajan, Amy Karger, and Sophie Arbefeville), the Department of Microbiology & Immunology (Drs. Ryan Langlois and Ashley Haase) and the Medical School Office of the Dean (Drs. Jakub Tolar and Timothy Schacker, Lisa Johnson, and staff). Finally, we want to thank the leadership and countless clinical laboratory staff from M Health Fairview including: Kylene Karnuth, Shannon Gascoigne, Michaela Leary, Jessica Gunderson, Klint Kjeldahl, Wendy Walters and many others.

