## Supplemental figures and tables for "Saliva testing is accurate for early-stage and presymptomatic COVID-19"

**AJ Johnson et al.**

**Table of Contents:**

Supplemental Figure 1: Results from N2 primers agreement with N1 primers. 2

Supplemental Figure 2: Relative viral load results. 3

Supplemental Figure 3: Evaluation of Cohort 1B symptoms, cycle thresholds, and test platform. 4

Supplemental Table 1: Contingency comparisons for Cohort 1A. 5

Supplemental Table 2: Contingency comparisons for Cohort 2. 5

Supplemental Table 3: Contingency comparisons for Cohort 1B. 5

Clinical Quality Assurance Data. 6

Supplemental Table 4: Contingency comparison of CLIA-LDT and Cepheid. 6

Supplemental Table 5: Contingency comparison of CLIA-LDT and Diasorin. 6

References. 6

**Supplemental Figure 1: Results from N2 primers agreement with N1 primers.**

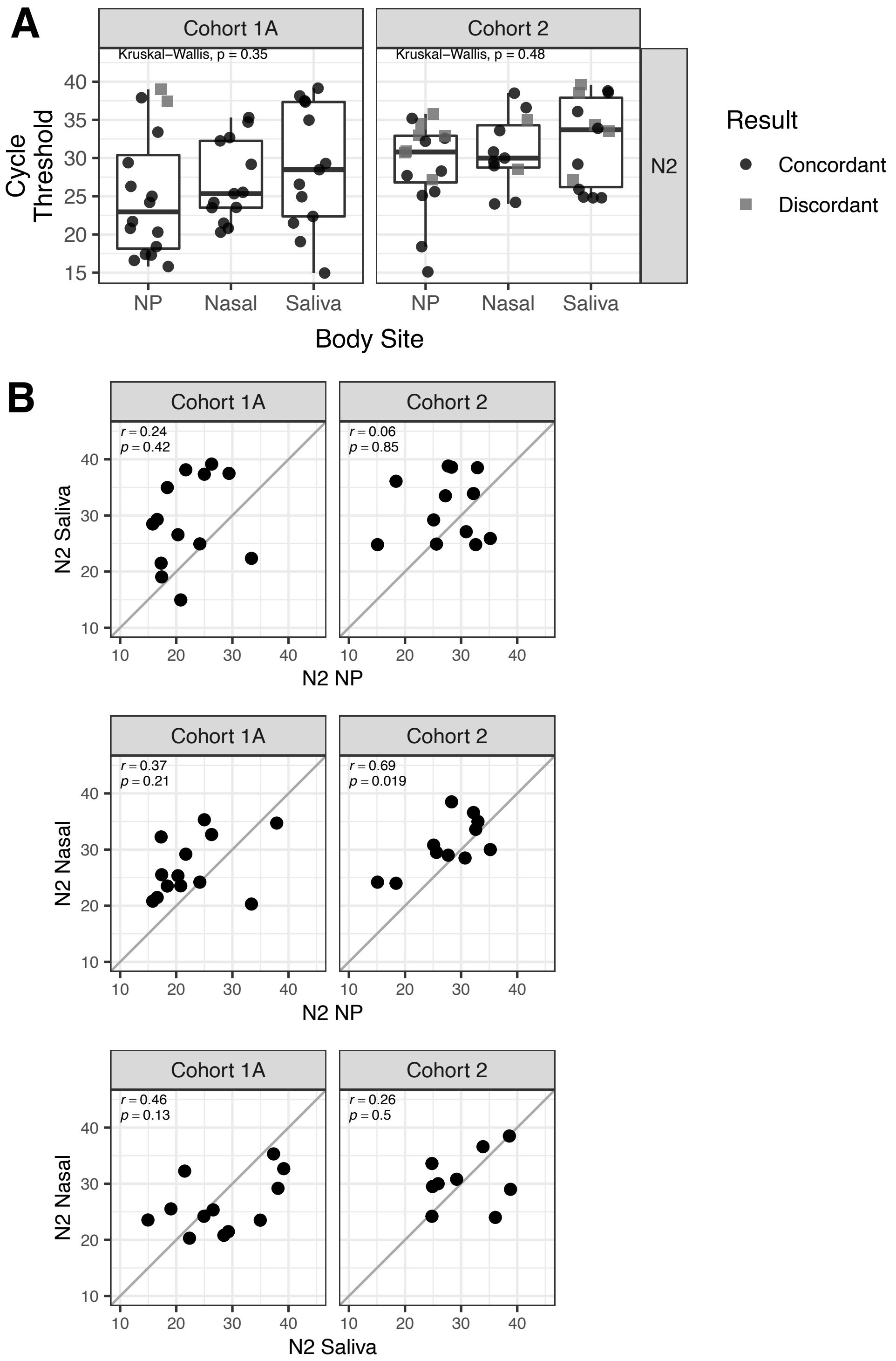

A) Box plots showing cycle threshold (Ct) values for Cohort 1A (n = 41) and 2 (n = 18) from the CLIA-LDT assay) for nasopharyngeal samples (NP), nasal samples, and saliva samples using N2 primers. Black circles are samples with concordance between nasal/saliva samples and NP samples (positive/positive or negative/negative). Grey squares show discordance between nasal/saliva samples and NP samples (positive/negative). Groups are compared using Kruskal-Wallis test by ranks. B) Scatter plots of Ct values from N2 primers for concordant samples from different sample types: saliva v. NP; nasal v. NP; nasal v. saliva. Correlation was using Pearson's correlation. The correlation coefficient (r) and p-value for each comparison are indicated on each plot.

**Supplemental Figure 2: Relative viral load results.**

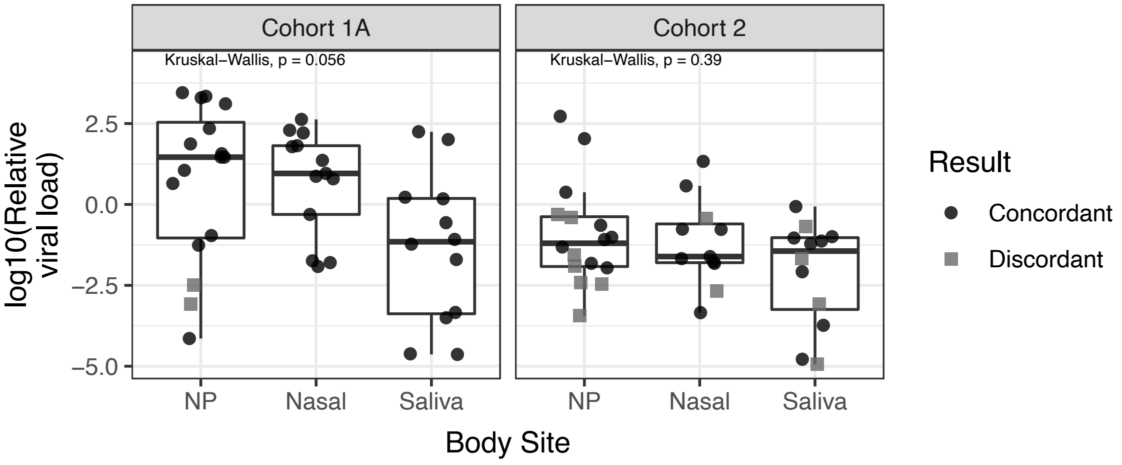

A) Box plots showing calculated relative viral load values for Cohort 1A (n = 41) and 2 (n = 18) for nasopharyngeal samples (NP), nasal samples, and saliva samples using N2 primers. Black circles are samples with concordance between nasal/saliva samples and NP samples (positive/positive or negative/negative). Grey squares show discordance between nasal/saliva samples and NP samples (positive/negative). Groups are compared using Kruskal-Wallis test by ranks.

**Supplemental Figure 3: Evaluation of Cohort 1B symptoms, cycle thresholds, and test platform.**

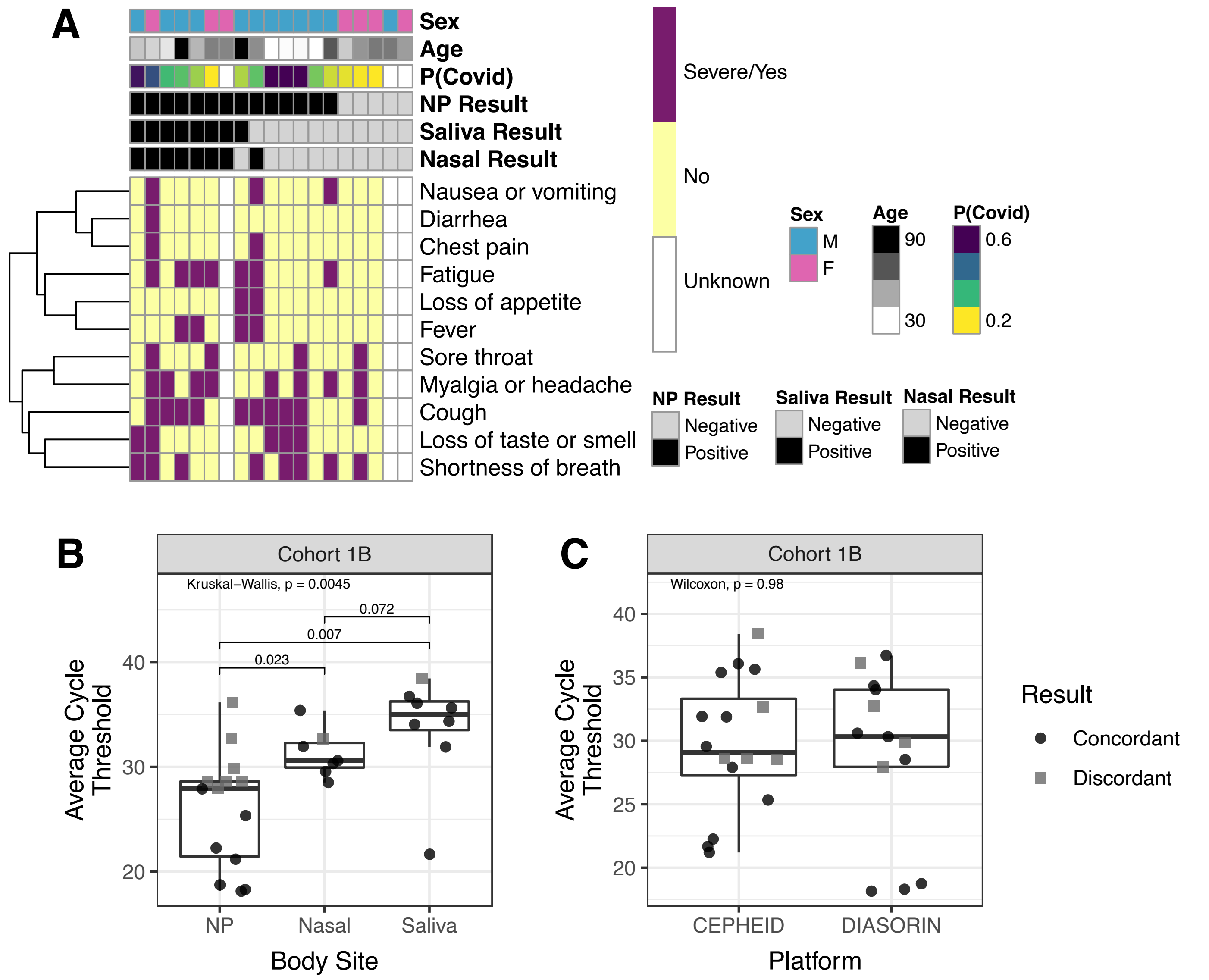

A) Heat map columns are participants and rows are symptoms. Symptom presence is indicated as either severe or yes (purple), no (yellow), or unknown (white). Symptoms are clustered by similarity and clustering is indicated by the dendrogram (left). Rows above the heatmap show metadata about each participant. Test results are indicated by color as either positive (black) or negative (light grey). Each participant's calculated probability of COVID-19 (P(COVID)) is indicated with higher probabilities shown in purple and lower probabilities in yellow. Participant sex, age, and cohort are also indicated. B) Box plots showing average cycle threshold (Ct) values (mean of N1 and N2 primers) for Cohort 1B (n = 19) for nasopharyngeal samples (NP), nasal samples, and saliva samples. Groups are compared using Kruskal-Wallis test by ranks. C) Box plots showing average Ct values (mean N1 and N2 primers) for Cohort 1B for each platform used. For all plots black circles are samples with concordance between nasal/saliva samples and NP samples (positive/positive or negative/negative). Grey squares show discordance between nasal/saliva samples and NP samples (positive/negative).

### Supplemental Table 1: Contingency comparisons for Cohort 1A.

| Cohort 1A | NP | | |
|---|---|---|---|
| Nasal | Positive | Negative | Total |
| Positive | 14 | 0 | 14 |
| Negative | 2 | 25 | 27 |

| PPA | OPA |
|---|---|
| 87.50% | 95.12% |

| Cohort 1A | NP | | |
|---|---|---|---|
| Saliva | Positive | Negative | Total |
| Positive | 14 | 0 | 14 |
| Negative | 2 | 25 | 27 |

| PPA | OPA |
|---|---|
| 87.50% | 95.12% |

| Cohort 1A | Nasal | | |
|---|---|---|---|
| Saliva | Positive | Negative | Total |
| Positive | 14 | 0 | 14 |
| Negative | 0 | 27 | 27 |

| PPA | OPA |
|---|---|
| 100.00% | 100.00% |

### Supplemental Table 2: Contingency comparisons for Cohort 2.

| Cohort 2 (n=20) | NP | | |
|---|---|---|---|
| Nasal | Positive | Negative | Total |
| Positive | 11 | 0 | 11 |
| Negative | 5 | 4 | 9 |

| PPA | OPA |
|---|---|
| 68.75% | 75.00% |

| Cohort 2 (n=20) | NP | | |
|---|---|---|---|
| Saliva | Positive | Negative | Total |
| Positive | 12 | 2 | 14 |
| Negative | 4 | 2 | 6 |

| PPA | OPA |
|---|---|
| 75.00% | 70.00% |

| Cohort 2 (n=20) | Nasal | | |
|---|---|---|---|
| Saliva | Positive | Negative | Total |
| Positive | 9 | 5 | 14 |
| Negative | 2 | 4 | 6 |

| PPA | OPA |
|---|---|
| 81.82% | 65.00% |

### Supplemental Table 3: Contingency comparisons for Cohort 1B.

| Cohort 1B | NP | | |
|---|---|---|---|
| Nasal | Positive | Negative | Total |
| Positive | 8 | 0 | 8 |
| Negative | 6 | 5 | 11 |

| PPA | OPA |
|---|---|
| 57.14% | 68.42% |

| Cohort 1B | NP | | |
|---|---|---|---|
| Saliva | Positive | Negative | Total |
| Positive | 8 | 0 | 8 |
| Negative | 6 | 5 | 11 |

| PPA | OPA |
|---|---|
| 57.14% | 68.42% |

| Cohort 1B | Nasal | | |
|---|---|---|---|
| Saliva | Positive | Negative | Total |
| Positive | 7 | 1 | 8 |
| Negative | 1 | 10 | 11 |

| PPA | OPA |
|---|---|
| 87.50% | 89.47% |

PPA: Positive Percent Agreement; OPA: Overall Percent Agreement

**Clinical Quality Assurance Data.**
The limit of detection of the CDC-based CLIA-LDT at our institution was validated between 560-1670 viral copies/mL depending on the specific viral extraction method used (4 distinct methods were validated to accommodate supply chain limitations). In comparison, the LOD for the commercial platforms were reported as 250 (Cepheid) and 500 (Diasorin) viral copies/mL in the respective EUA documents. Quality assurance (QA) data from our clinical laboratory shows the LDT has a PPA=90% (95% C.I. 74-96%) versus the Cepheid and a PPA=97% (95% C.I. 83-99%) vs. the Diasorin platforms in retrospective re-testing of archived samples on the LDT. All discordant samples in this QA data demonstrated late Ct values (>38) in the commercial test, consistent with variation around the limit of detection for these platforms. Furthermore, the LDT comparisons were inherently impaired, as all samples had been banked frozen and subjected to a freeze-thaw cycle prior to testing on the LDT. Parallel comparison was not possible to multiple factors, including: severe supply chain limitations on the commercial platforms prohibiting non-essential testing, and physical separation of the commercial and LDT platforms across the health care system that prevented simultaneous processing. The level of inter-platform agreement we have observed is aligned with the ranges published by others[1-3].

**Supplemental Table 4: Contingency comparison of CLIA-LDT and Cepheid.**

| | | Positive | Negative | | | |
|---|---|---|---|---|---|---|
| CLIA-LDT | Positive | 27 | 0 | | OPA | 93% |
| | Negative | 3 | 15 | | PPA | 90% |

**Supplemental Table 5: Contingency comparison of CLIA-LDT and Diasorin.**

| | | Diasorin | | | | |
|---|---|---|---|---|---|---|
| | | Positive | Negative | | | |
| CLIA-LDT | Positive | 29 | 0 | | OPA | 98% |
| | Negative | 1 | 15 | | PPA | 97% |

**References.**

1. Zhen W, Manji R, Smith E, Berry GJ. Comparison of four molecular in vitro diagnostic assays for the detection of sars-cov-2 in nasopharyngeal specimens. J Clin Microbiol. 2020;58(8).
2. Procop GW, Brock JE, Reineks EZ, et al. A comparison of five sars-cov-2 molecular assays with clinical correlations. Am J Clin Pathol. 2021;155(1):69-78.
3. Lieberman JA, Pepper G, Naccache SN, Huang M-L, Jerome KR, Greninger AL. Comparison of commercially available and laboratory-developed assays for in vitro detection of sars-cov-2 in clinical laboratories. J Clin Microbiol. 2020;58(8).
